# The association between clonal hematopoiesis driver mutations, immune cell function and the vasculometabolic complications of obesity

**DOI:** 10.1101/2023.07.07.23292396

**Authors:** Helin Tercan, Benjamin Cossins, Rosanne van Deuren, Joost H.W. Rutten, Prof. Leo A.B. Joosten, Prof. Mihai G. Netea, Alexander Hoischen, Siroon Bekkering, Prof. Niels P. Riksen

## Abstract

**Background:** Obesity is accompanied by dysregulated inflammation, which can contribute to vasculometabolic complications, including metabolic syndrome and atherosclerosis. Recently, clonal hematopoiesis of indeterminate potential (CHIP) has emerged as a risk factor for cardiovascular diseases. We aimed to determine how CHIP related to immune cell function, systemic inflammation, and vasculometabolic complications in obese individuals.

**Methods:** 297 individuals with overweight and obesity, between the ages of 54 and 81, were recruited in a cross-sectional study. Clonal Hematopoiesis Driver Mutations (CHDMs) were identified with an ultrasensitive targeted assay. Assesment of carotid artery atherosclerosis was performed with ultrasound. Detailed immunological parameters were studied. Adipose tissue inflammation was determined in subcutenous fat biopsies.

**Results:** Individuals with CHIP had higher concentrations of circulating IL-6. Total number of leukocytes and neutrophils were higher in individuals with CHIP. In contrast, *ex vivo* cytokine production capacity of PBMCs was significantly lower in individuals with CHIP. Sex stratified analysis showed that men with CHDMs had significantly higher leukocyte and neutrophil counts and *ex vivo* cytokine production capacity was lower in women with CHDMs. Surprisingly, the presence of atherosclerotic plaques was significantly lower in individuals with CHDMs. There was no relation between CHIP and metabolic syndrome.

**Conclusion:** In individuals with overweight or obesity, CHDMs are not associated with vasculometabolic complications, but rather with a lower presence of carotid plaques. CHDMs associate with increased circulating inflammatory markers and leukocyte numbers, but a lower PBMC cytokine production capacity.

## Introduction

The prevalence of overweight and obesity has increased globally in the past decades. This poses a great risk for cardiometabolic diseases, and a significant burden on the healthcare system^1^. Obesity is associated with the development of atherosclerosis, the leading cause of cardiovascular diseases (CVD), via metabolic dysregulation and chronic low-grade inflammation. Furthermore, obesity can induce activation of innate immune cells, such as monocytes, which are critical in the development of atherosclerotic CVD^2^.

Innate immune cell function is influenced by many factors, and one recently described mechanism is clonal hematopoiesis. Somatic mutations are common in highly proliferative tissues, including the hematopoietic stem and progenitor cells in the bone marrow. If, by chance, one of these mutations confers a selective survival or fitness advantage, it leads to the clonal expansion of that cell^3^. Recent advances in sequencing technologies demonstrated that clonal hematopoiesis is common among the aging population^4^. Clonal hematopoiesis driver mutations (CHDMs) are linked with an increased risk for hematological malignancies^5,6^, but multiple epidemiological studies also show that presence of CHDMs is associated with an increased risk of atherosclerotic CVD^7^.

Common somatically mutated genes include *DNMT3A (*DNA methyltransferase 3a), *TET2* (tet methylcytosine dioxygenase 2), *ASXL1* (ASXL transcriptional regulator 1), *JAK2* (Janus kinase 2) and *TP5*3 (tumor protein p53)^3^. Clonal hematopoiesis of indeterminate potential (CHIP) is defined as the presence of CHDMs with a variant allele frequency (VAF) ≥2% without clinical diagnosis of a hematological malignancy^5^. This cutoff is predominantly determined by the limitations of standard sequencing methods, including whole-exome sequencing, frequently used to identify CHDMs. Recently, the 2% cutoff has been challenged, underscoring the potential clinical relevance of driver mutations with lower VAFs^8^.

A few experimental pre-clinical studies suggest that CHDMs are involved in the metabolic and cardiovascular complications of obesity. For example, experimental studies in mice showed that TET2 driven clonal hematopoiesis is causal to age- and obesity-related insulin resistance. This is mediated by the activation of innate immune cells via NLRP3-inflammasome driven IL-1β production^9^. In the general population, the presence of CHDM is associated with higher waist-to-hip ratio^10^. There are also indications for reverse causality, with atherosclerosis-associated inflammation accelerating clonal hematopoiesis in experimental mouse models^11^. Using the exact same sequencing technique and gene panel as in our study, Andersson-Assarsson *et al.*, recently reported an increase in time in obese individuals, but not in those who underwent bariatric surgery^12^.

Here, we propose that clonal hematopoiesis is an important driver of inflammation in patients with obesity, predominantly associated with older age, and of the subsequent development of metabolic syndrome and atherosclerosis. We hypothesize that this is mediated by increased monocyte responsiveness. To this end, we identified a panel of candidate CHDMs with an ultrasensitive assay in a cohort of 297 human subjects with overweight and obesity. We associated CHDMs to parameters of metabolic and atherosclerotic complications, to parameters of systemic inflammation, as well as to the innate immune cell phenotype and function. Since the regulation of inflammation in relation to metabolic syndrome is highly sex-specific, we performed all analyses stratified by sex^13^.

## Methods

### Study subjects and clinical measurements

This cross-sectional single center cohort study was part of the Human Functional Genomics Project (<u>Human Functional Genomics Project</u>). A cohort of 302 individuals (“the 300 OB cohort”) with a BMI> 27 kg/m^2^, between the ages of 54 and 82 was recruited in the Radboud university medical center from 2014 to 2016. The research protocol was approved by the Radboud University Ethical Committee (nr. 46846.091.13), and all subjects gave written informed consent. The study protocol was performed in accordance with the 1975 Declaration of Helsinki.

For an extensive description of the cohort and measurements please see ter Horst *et al.*, 2020^13^.

Lipid lowering medication was stopped four weeks before measurements. Blood samples were drawn in the morning after an overnight fast. Total blood cell counts were performed in fresh EDTA blood samples using a Sysmex Hematoanalyzer XE5000. Plasma samples were stored at -80°C until further measurement. Blood glucose, triglycerides, total cholesterol, high-density lipoprotein cholesterol (HDL-C) and low-density lipoprotein cholesterol (LDL-C) were determined by standard laboratory protocols. Systolic and diastolic blood pressure were measured after 30 minutes of supine rest.

Metabolic syndrome was defined according to the National Cholesterol Education Program ATP III criteria^14^. Participants having at least 3 of the following characteristics were determined to have metabolic syndrome:

1. Abdominal obesity: Waist circumference ≥ 102cm in men, or ≥ 88cm in women.
2. Triglycerides: serum TG ≥ 150 mg/dL (1.7 mmol/L), or treatment with lipid-lowering drugs.
3. HDL Cholesterol: serum HDL-C < 40 mg/dL (1 mmol/L) in men, or < 50 mg/dL (1.3 mmol/L) in women.
4. Blood pressure: ≥ 130/85 mmHg, or treatment for hypertension.
5. Fasting plasma glucose: ≥ 100 mg/dL (5.6 mmol/L), or treatment for elevated blood glucose.

Insulin resistance was determined according to the HOMA model^15^.

### Cardiovascular measurements

Carotid artery measurements were performed by ultrasound, as described previously^16^. Carotid intima-medial thickness (IMT) was determined in the proximal 1 cm straight portion of the carotid artery at 90°, 120° and 180° angles for 6 heart beats. The presence and thickness of plaques were assessed in the common carotid, internal carotid, or external carotid artery or at the carotid bulbus. A focal thickening of the carotid wall of at least 1.5x IMT or an IMT>1.5 mm was termed as plaque presence. Pulse Wave Velocity (PWV) and Augmentation Index adjusted for heart rate (AI) were measured to assess arterial stiffness by SphygmoCor (ATCOR Medical) under standard operating protocol.

### Adipose tissue analysis

Detailed methodology regarding adipose tissue measurements were previously described^13^. Briefly, subcutaneous abdominal adipose tissue samples were obtained by needle biopsies under local anesthesia. Adipocyte size, presence of macrophages, and crown-like structures were determined by fluorescent microscopy (Zeiss Axiphot) utilizing Hematoxylin and Eosin (H&E) in combination with CD68 staining. Additionally, total RNA was isolated from adipose tissue samples by Trizol (Invitrogen) extraction, followed by cDNA library preparation (iScript cDNA synthesis kit, Bio-Rad). mRNA expression levels were determined by real-time PCR and normalized to housekeeping gene (Ribosomal Protein L37a, RPL37A) expression.

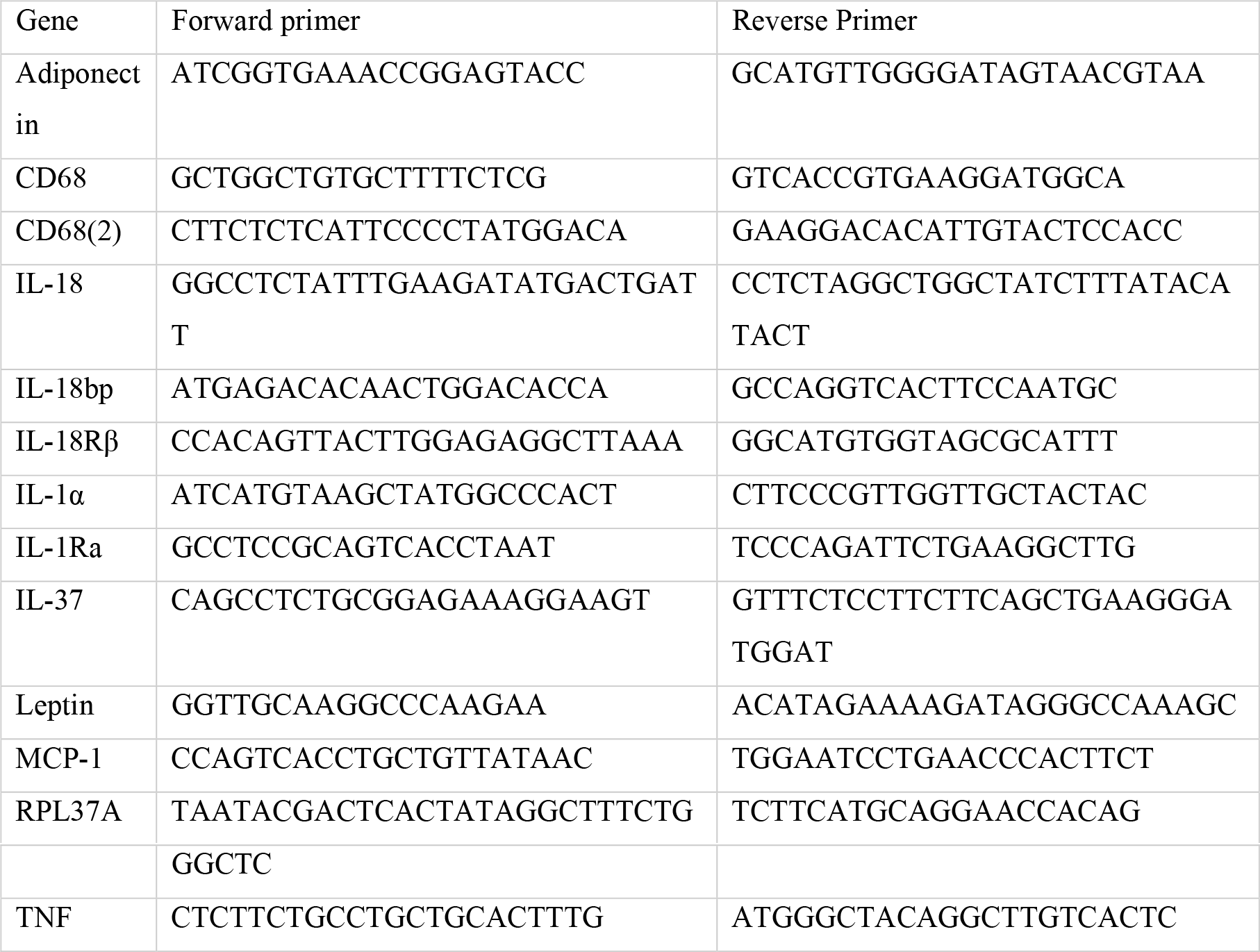

### Identification of CHDMs

CHDMs were identified in whole blood by an ultra-sensitive assay, as previously described^12,17^. In short, 300 single-molecule molecular inversion probes (smMIP) were designed against a selection of well-known hotspots of a panel of 24 clonal hematopoiesis driver genes (Supp. table 11). The *DNMT3A* gene, which contains the most driver mutations, was entirely covered^18^. We used the smMIP probe panel designed in 2017 ^19^, that was also used in recent studies^12,17^ (Supp. table 12). For each sample two technical PCR replicates were run, thereafter two independent data processing strategies and a quality control step were performed. Variant allele frequencies were calculated using samtools mpileup^20^ (Supp. table 13).

All identified CHDMs were validated in publically available datasets. For individuals with more than one CHDM, the CHDM with the highest VAF was included in the analysis. We divided all identified CHDMs into: “Low VAF” for CHDMs with a VAF<2% and “High VAF” for CHDMs with a VAF≥2% in accordance with the current CHIP cutoff.

From the 302 individuals of the cohort, whole blood samples of 3 individuals could not be obtained, and the samples from 2 individuals did not pass the quality control for sequencing. Therefore, 5 individuals were completely excluded from this study, corresponding to 297 individuals for the final analysis.

### PBMC isolation and *ex vivo* stimulation

Peripheral Blood Mononuclear Cells (PBMCs) were isolated with differential density centrifugation over Ficoll-Paque(GE Healthcare). PBMCs were then washed thrice by centrifugation with phosphate saline buffer. Isolated PBMCs were resuspended in Dutch modified Roswell Park Memorial Institute (RPMI) 1640 medium (Invitrogen) supplemented with 50 µg/mL gentamicin (Centrafarm), 2 mM GlutaMAX and 1 mM pyruvate (Life Technologies). 0.5×10^6^ cells per well were stimulated for 24 hours in 96-wells round-bottom plates (Greiner) at 37 °C and 5% CO_2_. Stimuli used include RPMI as negative control, Lipopolysaccharide (LPS) (Sigma-Aldrich, E. coli serotype 055:B5, further purified as described^21^) at low (1 ng/ml) and high concentrations (100 ng/ml) and Pam3Cys (1 ug/ml) (EMC microcollections, L2000). Supernatants were collected after 24 hours and stored at -20°C until measurements were performed.

### Cytokine measurements

Cytokine concentrations upon stimulation of PBMCs and in plasma were measured with commercially available Enzyme-linked Immunosorbent Assay (ELISA) kits according to instructions supplied by the manufacturer.

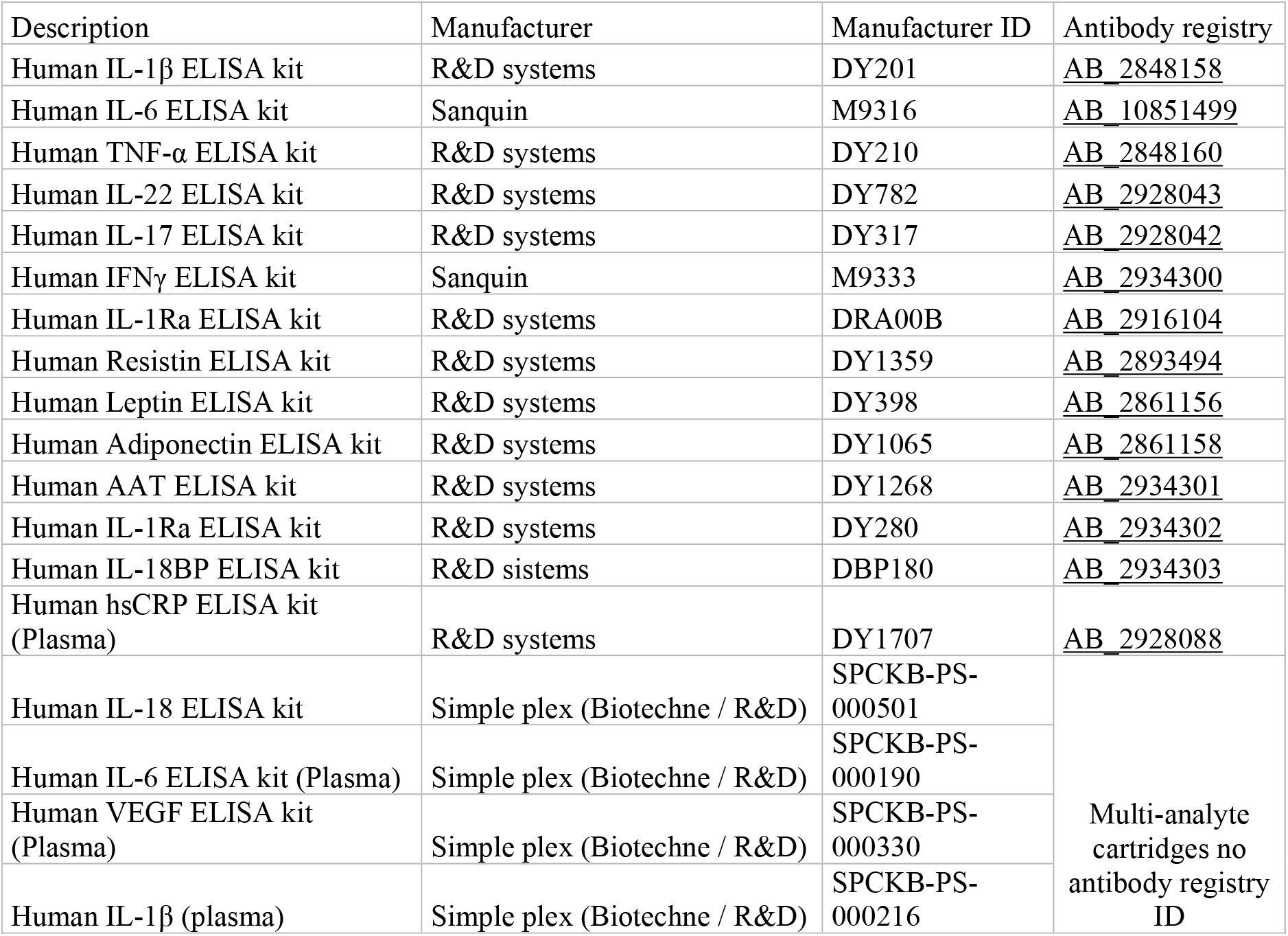

| Description | Manufacturer | Manufacturer ID | Antibody registry |
| --- | --- | --- | --- |
| Human IL-1β ELISA kit | R&D systems | DY201 | AB 2848158 |
| Human IL-6 ELISA kit | Sanquin | M9316 | AB 10851499 |
| Human TNF-α ELISA kit | R&D systems | DY210 | AB 2848160 |
| Human IL-22 ELISA kit | R&D systems | DY782 | AB 2928043 |
| Human IL-17 ELISA kit | R&D systems | DY317 | AB 2928042 |
| Human IFNγ ELISA kit | Sanquin | M9333 | AB 2934300 |
| Human IL-1Ra ELISA kit | R&D systems | DRA00B | AB 2916104 |
| Human Resistin ELISA kit | R&D systems | DY1359 | AB 2893494 |
| Human Leptin ELISA kit | R&D systems | DY398 | AB 2861156 |
| Human Adiponectin ELISA kit | R&D systems | DY1065 | AB 2861158 |
| Human AAT ELISA kit | R&D systems | DY1268 | AB 2934301 |
| Human IL-1Ra ELISA kit | R&D systems | DY280 | AB 2934302 |
| Human IL-18BP ELISA kit | R&D systems | DBP180 | AB 2934303 |
| Human hsCRP ELISA kit (Plasma) | R&D systems | DY1707 | AB 2928088 |
| Human IL-18 ELISA kit | Simple plex (Biotechne / R&D) | SPCKB-PS-000501 | Multi-analyte cartridges no antibody registry ID |
| Human IL-6 ELISA kit (Plasma) | Simple plex (Biotechne / R&D) | SPCKB-PS-000190 |  |
| Human VEGF ELISA kit (Plasma) | Simple plex (Biotechne / R&D) | SPCKB-PS-000330 |  |
| Human IL-1β (plasma) | Simple plex (Biotechne / R&D) | SPCKB-PS-000216 | |

### Proteomic profiling

The concentrations of 177 inflammatory proteins from the Olink Cardiovascular II and Inflammatory panels were measured using the previously described proximity extension assays (PEA) technology (Olink Bioscience AB, Uppsala, Sweden)^22^. Quality control was ensured using internal extension control and inter-plate control. The data is presented in Normalized Protein eXpression (NPX) values, an arbitrary unit on log2 scale.

### Statistics

Distribution of data was assessed with Shapiro-Wilk test. Normally distributed data is shown as mean ± standard deviation. If the data did not follow a normal distribution, it is shown as median and interquartile ranges. Each group (All CHDM, High VAF, and Low VAF) was independently compared to the subjects without CHDM (‘No CHDM group’) with Student’s t-test or Mann-Whitney U test when appropriate. Spearman correlation was used to determine the association between VAF and various clinical and immunological parameters. The same statistical methodology was applied for the sex specific analyses. P<0.05 is considered statistically significant and is indicated with an asterisk in tables. All statistical analyses were performed in R version 4.1.1 (R Core Team). Part of the data is publically available on the website of Human Functional Genomics Project (<u>HFGP (bbmri.nl)</u>), the rest is available upon reasonable request to the HFGP committee.

## Results

The study population included 297 individuals with a median age of 67 years (interquartile 1 and 3, 63-71 years) and BMI of 30 kg/m^2^ (28-32 kg/m^2^). Men represented 55% of the cohort. 54% of the cohort met the NCEP ATP III criteria of Metabolic Syndrome, and approximately half of the participants had carotid atherosclerotic plaques as measured by ultrasound. Hypertension was diagnosed in 60% of the study population, while diabetes was present in 12%. Median total cholesterol concentration was 6.3 mmol/l and triglyceride concentration was 1.61 mmol/l. 27% of the cohort used lipid lowering drugs. Detailed baseline characteristics of the study population is presented in Table 1.

**Table 1:**
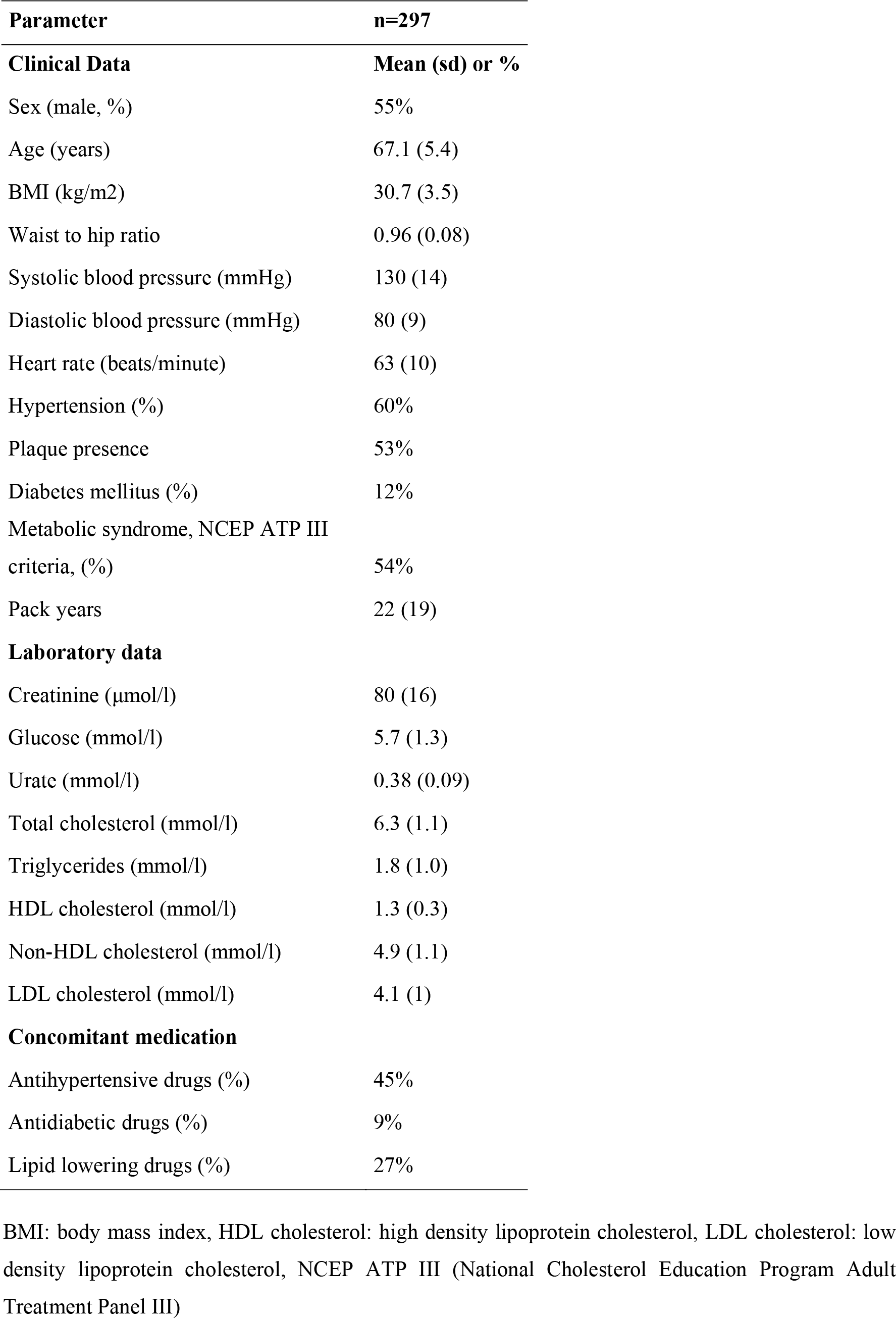
Baseline characteristics of the study cohort.

| Parameter | n=297 |
| --- | --- |
| **Clinical Data** | **Mean (sd) or %** |
| Sex (male, %) | 55% |
| Age (years) | 67.1 (5.4) |
| BMI (kg/m<sup>2</sup>) | 30.7 (3.5) |
| Waist to hip ratio | 0.96 (0.08) |
| Systolic blood pressure (mmHg) | 130 (14) |
| Diastolic blood pressure (mmHg) | 80 (9) |
| Heart rate (beats/minute) | 63 (10) |
| Hypertension (%) | 60% |
| Plaque presence | 53% |
| Diabetes mellitus (%) | 12% |
| Metabolic syndrome, NCEP ATP III criteria, (%) | 54% |
| Pack years | 22 (19) |
| **Laboratory data** |  |
| Creatinine (μmol/l) | 80 (16) |
| Glucose (mmol/l) | 5.7 (1.3) |
| Urate (mmol/l) | 0.38 (0.09) |
| Total cholesterol (mmol/l) | 6.3 (1.1) |
| Triglycerides (mmol/l) | 1.8 (1.0) |
| HDL cholesterol (mmol/l) | 1.3 (0.3) |
| Non-HDL cholesterol (mmol/l) | 4.9 (1.1) |
| LDL cholesterol (mmol/l) | 4.1 (1) |
| **Concomitant medication** |  |
| Antihypertensive drugs (%) | 45% |
| Antidiabetic drugs (%) | 9% |
| Lipid lowering drugs (%) | 27% |

BMI: body mass index, HDL cholesterol: high density lipoprotein cholesterol, LDL cholesterol: low density lipoprotein cholesterol, NCEP ATP III (National Cholesterol Education Program Adult Treatment Panel III)

### CHDM prevalence and characteristics

110 candidate CHDMs were identified in 85 individuals; 62 individuals (21% of the cohort) carried a single CHDM and 23 individuals (8%) had more than one CHDM (Fig 1A). The VAF of all CHDMs ranged from 0.01% to 34.5%, with a mean of 3.3% and median of 1.1%.

**Figure 1:**
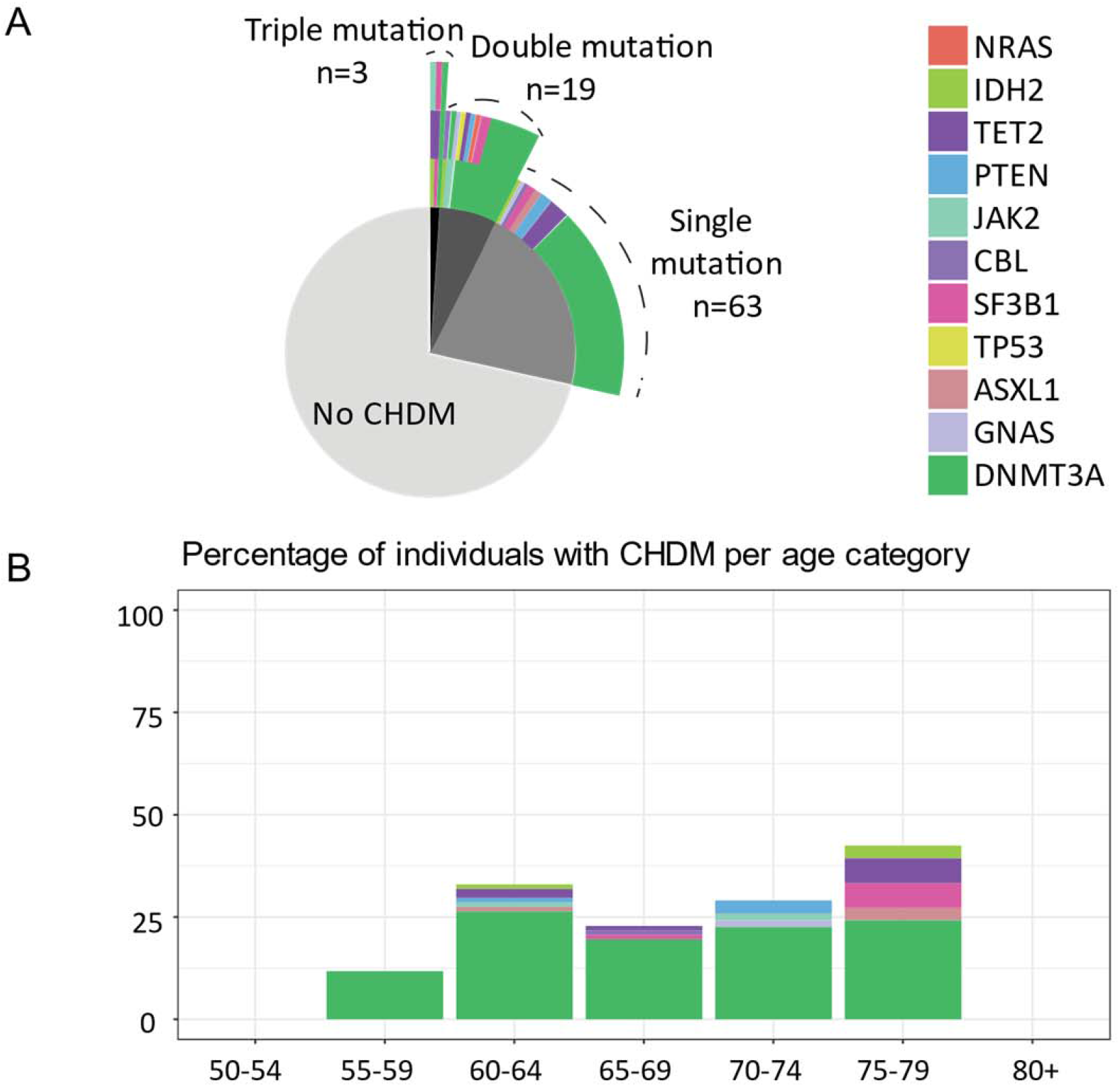
Characterization of candidate CHDMs identified in our study. A) Pie chart indicating number of individuals without CHDMs, with a single mutation, double mutations and triple mutations. Top rings indicate genes affected from the mutations. For more than one mutation carrying individuals second and third mutations are displayed as multiple rings on top of each other. B) Percentage of individuals with CHDMs per age category and gene affected.

In 33 individuals, we identified CHIP (i.e. CHDM with a VAF≥2%), and 52 individuals had CHDMs with VAF<2%. We identified CHDMs in 11 individual genes. Mutations in *DNMT3A* (73%) and *TET2* (7%) genes were the most common in the entire cohort (Fig 1A). Additionally, *DNMT3A* mutations were the most common across all age groups (Fig 1B).

### Relation between CHDM and clinical characteristics

We did not observe statistically significant differences in baseline characteristics between subjects with and without CHDMs, apart from the finding that individuals with CHDMs with VAF<2% presented with higher heart rate (Table 2). Although there was no significant association between sex and presence of CHDMs, we observed a trend towards more women in the All CHDM group (p=0.054). Sex-specific analysis revealed that men with CHIP were significantly older than those without CHDMs and men with CHDMs with VAF<2% had significantly higher heart rate (Supp. table 1). In women, there were no differences in baseline characteristics in any of the groups (Supp. table 2). We did not observe a correlation between presence or size of the clones and any of the other baseline parameters.

**Table 2:** Baseline characteristics according to CHDM status.

|  | **No CHDM**<br>**(n=212)** | **All CHDM**<br>**(n=85)** | **High VAF**<br>**(n=33)** | **Low VAF**<br>**(n=52)** | **Correlation**<br>**with VAF** |
| --- | --- | --- | --- | --- | --- |
| Age (years) | 66 (63-70) | 68 (63-72) | 67 (63-74) | 68 (63-71) | 0,1 |
| Sex (% Male) | 58 | 46 | 45 | 46 |  |
| Weight (kg) | 88 (82-97) | 89 (81-99) | 89 (80-98) | 87 (82-99) | -0,1 |
| Height (cm) | 172 (165-178) | 171 (164-176) | 172 (164-176) | 171 (164-177) | -0,1 |
| BMI (kg/m<sup>2</sup>) | 30 (28-32) | 30 (28-32) | 30 (28-32) | 30 (28-32) | 0 |
| Creatinine (μmol/L) | 81 (67-92) | 77 (68-89) | 76 (70-88) | 78 (67-89) | -0,2 |
| Glucose (mmol/L) | 5.4 (5-6) | 5.4 (5-6) | 5 (5-6) | 5.4 (5-6) | 0,004 |
| Total cholesterol (mmol/L) | 6 (6-7) | 6 (5-7) | 6 (5-7) | 6 (6-7) | -0,1 |
| Triglycerides (mmol/L) | 1.6 (1.3-2.2) | 1.5 (1.3-2) | 1.5 (1.3-2) | 1.5 (1.3-2) | -0,023 |
| Heart rate (beats/min) | 61 (55-67) | 64 (58-71) | 62 (59-67) | 66 (57-73) * | -0,2 |
| Antihypertensives (%) | 47 | 40 | 52 | 33 |  |
| Lipid lowering drugs (%) | 28 | 25 | 18 | 29 |  |
| Antidiabetic drugs (%) | 9 | 7 | 6 | 8 |  |

All data are given as median (interquartile ranges 1-3). \*indicates p<0.05 compared to No CHDM group. Correlation is indicated with Spearman correlation coefficient.

### Association of CHDM with parameters of metabolic dysregulation

We investigated the association between presence of CHDMs and presence of metabolic syndrome and its individual components, and parameters related to insulin resistance and diabetes mellitus type 2. We did not observe a higher prevalence of metabolic syndrome, its individual components or diabetes in individuals with clonal hematopoiesis (Table 3).

**Table 3:** Parameters of metabolic dysregulation (metabolic syndrome and diabetes) according to CHDM status.

|  | No CHDM<br>(n=212) | All CHDM<br>(n=85) | High VAF<br>(n=33) | Low VAF<br>(n=52) | Correlation<br>with VAF |
| --- | --- | --- | --- | --- | --- |
| Diabetes mellitus(%) | 14 | 9 | 9 | 10 |  |
| Metabolic syndrome (%) | 55 | 52 | 55 | 50 |  |
| Liver fat (mg/cm<sup>3</sup>) | 0.058 (0.024-0.12) | 0.062 (0.029-0.16) | 0.059 (0.034-0.14) | 0.066 (0.017-0.17) | -0,01 |
| Waist circumference (cm) | 106 (100-110) | 105 (100-112) | 106 (100-110) | 104 (100-113) | -0,09 |
| Hip circumference (cm) | 110 (106-114) | 111 (107-114) | 109 (106-114) | 111 (109-114) | -0,09 |
| HDL cholesterol (mmol/l) | 1.27 (1.1-1.51) | 1.32 (1.14-1.51) | 1.32 (1.11-1.51) | 1.33 (1.14-1.52) | 0,02 |
| LDL cholesterol (mmol/l) | 4.15 (3.57-4.72) | 4.06 (3.29-4.64) | 4.04 (3.18-4.58) | 4.07 (3.36-4.81) | -0,07 |
| Systolic blood pressure (mmHg) | 129 (118-138) | 130 (123-140) | 129 (123-140) | 131 (123-140) | -0,05 |
| Diastolic blood pressure (mmHg) | 80 (74-85) | 80 (72-87) | 76 (72-87) | 80 (72-87) | -0,08 |
| HOMA-IR | 7 (4-11) | 7 (4-16) | 5 (4-12) | 8 (4-16) | -0,13 |

HOMA-IR: Homeostatic Model Assessment of Insulin resistance, mmHg: millimeter of mercury
All data are given as median (interquartile ranges 1-3). \*indicates p<0.05 compared to No CHDM group. Correlation is indicated with Spearman correlation coefficient.

### Association between adipose tissue inflammation and presence of CHDMs

Adipose tissue is an important site for generation of various cytokines and adipokines, and is strongly associated with the development of cardiometabolic complications of obesity. Therefore, we characterized adipose tissue biopsies by immunohistochemistry and qPCR and explored whether adipose tissue inflammation is more severe in individuals with CHDMs.

Although we did not identify higher number of CD68^+^ macrophages by immunohistochemistry, we observed significantly higher expression of CD68, Adiponectin and TNF in individuals with CHDMs with VAF<2% measured with qPCR (Supp. Table 3).

### Relationship between CHDM and parameters of atherosclerosis

To explore whether the presence of CHDM correlates with the presence of atherosclerosis in subjects with obesity, we investigated IMT and carotid plaque presence and characteristics in relation to CHDM. We found that the presence of carotid plaques was lower in individuals with any CHDM and CHDMs with VAF<2% compared to individuals without CHDMs. Individuals with CHIP mutations had a significantly higher augmentation index (AI) suggestive of an increased systemic arterial stiffness (Table 4).

**Table 4:** Carotid IMT (cIMT) and parameters of carotid plaques and measures of arterial stiffness according to CHDM status of the participants.

|  | **No CHDM**<br>**(n=212)** | **All CHDM**<br>**(n=85)** | **High VAF**<br>**(n=33)** | **Low VAF**<br>**(n=52)** | **Correlation**<br>**with VAF** |
| --- | --- | --- | --- | --- | --- |
| cIMT ($\mu\text{m}$) | 791(703-884) | 753(691-859) | 815(713-882) | 745(692-832) | 0,11 |
| Carotid plaque presence (%) | 59 | 38 * | 48 | 31 * |  |
| Number of plaques | 1(1-2) | 1(1-2) | 2(1-2) | 1(1-2) | 0,16 |
| Max plaque thickness (mm ) | 2.2 (1.8-2.8) | 2.65 (1.98-3.2) | 2.75 (2-3.2) | 2.55 (1.87-2.97) | 0,17 |
| Max stenosis (%) | 0.29 (0.23-0.39) | 0.3 (0.25-0.39) | 0.37 (0.27-0.4) | 0.29 (0.22-0.34) | 0,25 |
| PWV (m/s) | 9.2 (8.4-10.7) | 9.4 (8.1-10.4) | 9.9 (8.2-10.4) | 9.3 (8.1-10.9) | -0,03 |
| Augmentation index (%) | 25.7 (20.5-30.3) | 27.5 (21.6-33.3) | 29.9 (24.6-34.8)* | 25.3 (19-32.1) | 0,31 |
| History of CVD (%) | 16 | 14 | 15 | 13 |  |

PWV: Pulse wave velocity, PWA: Pulse wave analysis, CVD: Cardiovascular disease
All data are given as median (interquartile ranges 1-3). \*indicates $p < 0.05$ compared to No CHDM group. Correlation is indicated with Spearman correlation coefficient.

### Leukocyte number and function and presence of CHDMs

The absolute number of leukocytes and thrombocytes were measured in whole blood. We identified a significantly higher number of total leukocytes, and specifically neutrophils, in individuals with CHDMs and in CHIP carriers. There were no differences in monocyte or lymphocyte counts (Table 5). Sex-specific analysis revealed men with CHIP had significantly higher neutrophil-to-lymphocyte ratio (Supp. Table 4). In contrast, there was no difference in any leukocyte count in women (Supp. Table 5).

**Table 5:** Leukocyte numbers and differentiation, and thrombocyte numbers separated according to CHDM status of the participants.

|  | **No CHDM**<br>**(n=212)** | **All CHDM**<br>**(n=85)** | **High VAF (n=33)** | **Low VAF (n=52)** | **Correlation**<br>**with VAF** |
| --- | --- | --- | --- | --- | --- |
| Leukocytes $10^9/\text{l}$ | 5.7 (5-6.6) | 6.1 (5.3-7.4) * | 6.2 (5.5-7.6) * | 5.9 (5.3-7.4) | 0,15 |
| Neutrophils $10^9/\text{l}$ | 3.2 (2.6-3.7) | 3.4 (2.7-4.4) * | 3.6 (2.9-4.5) * | 3.2 (2.7-4.3) | 0,13 |
| Lymphocytes $10^9/\text{l}$ | 1.8 (1.5-2.2) | 1.9 (1.6-2.3) | 1.9 (1.6-2.2) | 1.9 (1.6-2.3) | 0,1 |
| Monocytes $10^9/\text{l}$ | 0.5 (0.4-0.6) | 0.5 (0.4-0.6) | 0.5 (0.4-0.6) | 0.5 (0.4-0.6) | 0,1 |
| Eosinophils $10^9/\text{l}$ | 0.16 (0.09-0.21) | 0.15 (0.1-0.29) | 0.13 (0.09-0.29) | 0.16 (0.11-0.22) | 0,002 |
| Basophils $10^9/\text{l}$ | 0.03 (0.02-0.04) | 0.03 (0.02-0.04) | 0.03 (0.02-0.05) | 0.03 (0.02-0.04) | -0,03 |
| Thrombocytes $10^9/\text{l}$ | 225 (194-262) | 231 (196-266) | 226 (203-281) | 231(196-264) | 0,18 |
| NLR | 1.72 (1.31-2.18) | 1.8 (1.41-2.25) | 1.81 (1.5-2.42) | 1.78 (1.32-2.08) | 0,021 |

NLR: Neutrophil to lymphocyte ratio
All data are given as median (interquartile ranges 1-3). \*indicates $p < 0.05$ compared to No CHDM group. Correlation is indicated with Spearman correlation coefficient.

*Ex vivo* cytokine production capacity of PBMCs can be used as a measure of inflammatory responsiveness, and this has been shown to be higher in patients with established coronary heart disease^23,24^. Therefore, we characterized proinflammatory cytokine production capacity of PBMCs upon stimulation with Pam3Cys and LPS at different concentrations. Interestingly, we observed significantly lower production of IL-1β upon Pam3Cys stimulation in individuals with all CHDMs and CHDMs with VAF<2. Likewise, stimulation with LPS (both concentrations) led to significantly lower production of IL-6 in individuals with CHDMs, and CHIP carriers had significantly lower IL-6 upon stimulation with high concentration of LPS. Lastly, stimulation with Pam3Cys resulted in significantly less IL-6 production in all groups compared to individuals without any CHDMs (Table 6). Interestingly, the lower production of these cytokines in individuals with CHDMs was solely seen in women and not in men (Supp. tables 6 and 7).

**Table 6:** *Ex vivo* cytokine production capacity of PBMCs separated according to CHDM status of the participants.

|  | No CHDM<br>(n=212) | All CHDM<br>(n=85) | High VAF (n=33) | Low VAF (n=52) | Correlation<br>with VAF |
| --- | --- | --- | --- | --- | --- |
| LPS(1 ng/ml) IL-1β | 1075<br>(599-1936) | 998<br>(564-1710) | 1036<br>(564-2013) | 905<br>(561-1399) | 0,05 |
| LPS(100 ng/ml) IL-1β | 2476<br>(1544-3713) | 2210<br>(1245-3375) | 2703<br>(1280-3274) | 1823<br>(1240-3396) | 0,01 |
| Pam3Cys(1 μg/ml) IL-1β | 280<br>(111-640) | 174<br>(77-474) * | 163<br>(53-569) | 180<br>(82-424) * | -0,004 |
| LPS (1 ng/ml) IL-6 | 5750<br>(3278-8968) | 4850<br>(2969-6547) * | 4464<br>(2989-6547) | 5318<br>(2963-6449) | -0,06 |
| LPS (100 ng/ml) IL-6 | 8322<br>(5206-12536) | 7167<br>(4279-10797) * | 6062<br>(3606-9402) * | 7410<br>(4611-11114) | -0,09 |
| Pam3Cys (1 μg/ml) IL-6 | 3975<br>(1759-6545) | 2010<br>(1021-4548) * | 2010<br>(811-4548) * | 2034<br>(1364-4627) * | -0,05 |

LPS: Lipopolysaccharide, IL: interleukin, IL-1β and IL-6 concentration units are given in pg/ml
All data are given as median (interquartile ranges 1-3). \*indicates $p < 0.05$ compared to No CHDM group. Correlation is indicated with Spearman correlation coefficient.

### The relation between CHDMs and circulating cytokines and adipokines

To determine the association between systemic inflammation and presence of CHDMs we measured a selection of circulating cytokines and adipokines in plasma. We observed significantly higher concentrations of circulating IL-6 in individuals with CHIP. We identified a trend towards higher circulating IL-1β concentration in individuals with CHDMs with VAF<2%, although this did not reach statistical significance (p=0.053). We did not observe statistically significant differences in hsCRP concentrations. Additionally, individuals with CHDMs with VAF<2% had significantly higher concentrations of resistin in circulation (Table 7). We did not observe associations for any other circulating marker. There was no sex specific association between circulating markers and the presence of CHDMs (Supp. tables 8 and 9).

**Table 7:** Circulating cytokine and adipokine concentrations separated according to CHDM status of the participants.

|  | No CHDM<br>(n=212) | All CHDM (n=85) | High VAF (n=33) | Low VAF (n=52) | Correlation<br>with VAF |
| --- | --- | --- | --- | --- | --- |
| IL-6 (pg/ml) | 2.4(1.7-3.4) | 2.6(1.7-4.1) | 2.9(1.9-4.4) * | 2.4(1.6-3.4) | 0,18 |
| IL-1β (pg/ml) | 0.06 (0.06-0.11) | 0.06 (0.06-0.12) | 0.06 (0.06-0.13) | 0.07 (0.06-0.11)^ | -0,048 |
| IL-18 (pg/ml) | 304 (227-490) | 288 (216-484) | 320 (221-455) | 285 (210-505) | 0,024 |
| IL-18bp (ng/ml) | 17.2 (14.1-21) | 16.1 (13.3-19.2) | 16 (13-17.8) | 16.2 (14.1-20) | -0,14 |
| hsCRP (μg/ml) | 1.9 (0.9-3.3) | 2 (1.2-3.3) | 1.9 (0.9-3.9) | 2 (1.4-3.3) | 0,083 |
| AAT (mg/ml) | 0.9 (0.6-1.7) | 0.9 (0.6-1.4) | 0.9 (0.6-1.4) | 0.9 (0.6-1.4) | 0,019 |
| Resistin (ng/ml) | 10.6 (8.2-13.5) | 11.5 (9.2-15) | 11.6 (8.7-13.1) | 11.4 (9.5-16.2) * | -0,095 |
| Leptin (ng/ml) | 15.8 (9.7-29.5) | 18.1 (11.1-35.5) | 17.2 (9.3-25.5) | 19.1 (11.6-36.6) | -0,028 |
| Adiponectin<br>(μg/ml) | 4.3 (2.8-6) | 4.3 (3.1-5.7) | 4.3 (3.4-5.2) | 4.3 (3.1-6.4) | 0,15 |

IL: interleukin, IL-18bp: IL-18 binding protein, CRP: C-reactive protein, AAT: Alpha-1 antitrypsin,
^: p= 0.053
All data are given as median (interquartile ranges 1-3). \*indicates p<0.05 compared to No CHDM group. Correlation is indicated with Spearman correlation coefficient.

To further investigate the association between circulating proteins and CHDM status, we performed a targeted proteomics approach with Olink panels Cardiovascular II and Inflammation. Supplementary table 10 shows the list of proteins that were significantly different in individuals with CHDMs. However, these differences were not present after correction for multiple testing.

## Discussion

In this cross-sectional study of older overweight and obese individuals, we investigated the presence of clonal hematopoiesis driver mutations in relation to a wide range of clinical and immunological parameters. We hypothesized that the presence of CHDMs predisposes to the development of metabolic syndrome and atherosclerosis, and that this is mediated by immune cell activation and systemic inflammation. Approximately 28% of the individuals had a CHDM. We showed that individuals with CHDMs had higher numbers of leukocytes and neutrophils, and a higher circulating IL-6 concentration. In contrast, the *ex vivo* cytokine production capacity of PBMCs from individuals with CHDMs appeared to be lower compared to those without. On a clinical level, the presence of CHDMs of CHIP did not associate with the presence of metabolic syndrome or atherosclerosis. Rather, the presence of carotid plaques was significantly lower in individuals with CHDMs. Lastly, we identified that several of the associations with CHDMs were sex specific.

Acquisition of somatic mutations is a hallmark of aging, and obesity is known to accelerate the pace of aging^25^. Systemic inflammation and activation of the immune system drives the development of metabolic and atherosclerotic complications in obesity^26^. Thus, clonal hematopoiesis may be pivotal in the association between obesity and atherosclerotic cardiovascular disease in aging individuals.

Several epidemiological studies identified the most common clonal hematopoiesis driver mutations to be in *DNMT3A* and *TET2* genes^4,7^. While a VAF≥2% has been used traditionally to distinguish CHIP, recent advances in sequencing methodologies with superior sequencing depth have revealed that also smaller clone sizes can be associated with cardiovascular disease^27^. Recent work by Assmus *et al.* argued for mutation-specific cutoff values, 1.15% for DNMT3A and 0.73% for TET2, to predict all-cause mortality in heart failure patients^8^. Given the relevance of clones with smaller sizes, we included them in our analysis.

We observed a trend towards more women having CHDMs in this population. This finding prompted us to perform a sex-specific analysis. Men with CHIP were significantly older than those without CHDMs. However, women with or without CHIP were the same age. As women on average live longer than men, the clone growth might be delayed. Additionally, it has been shown that men generally have shorter telomere lengths, a trait associated with clonal hematopoiesis^28,29^.

We hypothesized that in obese individuals, CHDMs would predispose to metabolic syndrome and to atherosclerosis due to increased monocytes responsiveness and increased systemic inflammation. We observed higher leukocyte and neutrophil numbers in individuals with CHDMs, in particular when the VAF was ≥2% (i.e. in the presence of CHIP). In addition, the circulating IL-6 concentration was higher in individuals with CHIP, confirming previous findings^30^. A previous genetic study showed that the increased CVD risk associated with CHIP is abrogated in individuals with a genetically determined reduced IL-6 signaling^31^, although this was not confirmed in a larger study^18^. In contrast to the higher circulating IL-6, we did not find a higher cytokine production capacity in PBMCs of individuals with CHDMs; we rather observed a lower production capacity for IL-6 and IL-1β in women with CHDMs. This contrasts the experimental finding that TET2-deficient mouse macrophages had increased cytokine production capacity^32^. Furthermore, single cell RNA sequencing of unstimulated human monocytes from patients with heart failure revealed increased expression of inflammatory genes in individuals harboring *DNMT3A* mutations^27^. These discrepant findings might indicate a differential effect of CHDMs on the inflammatory phenotype of monocytes versus macrophages or on baseline inflammatory gene expression versus protein production upon stimulation. One potential explanation is that the circulating PBMCs might have developed tolerance by the continuous exposure to higher concentrations of cytokines, thus impairing the *ex vivo* cytokine production capacity upon stimulation. Interestingly, the lower cytokine production capacity in the presence of CHDMs is exclusively observed in women in the sex stratified analysis.

In contrast to our hypothesis, we did not observe more atherosclerosis in individuals with CHDMs, as assessed with carotid ultrasound measuring to IMT and atherosclerotic plaques. Surprisingly, the presence of carotid plaques was even significantly lower in these individuals. Of note, the effect of CHDMs or CHIP on non-symptomatic atherosclerotic plaques has never been studied before. Various studies showed a strong association of CHIP with the occurrence of cardiovascular events, which are mostly triggered by destabilization of atherosclerotic plaques and subsequent thrombus formation^4,8^. Thus, our data suggest that clonal hematopoiesis does not facilitate the initial development of atherosclerotic plaques, but rather the destabilisation of these plaques. In line with this, atherosclerotic plaque development starts as early as in the second or third decade of life while clonal hematopoiesis is primarily seen in individuals older than 55^5,33^.

Finally, there was no association between CHDMs or CHIP and markers of metabolic complications of obesity, including metabolic syndrome and its individual components, liver fat, and insulin resistance. Similarly, we did not observe increased adipose tissue inflammation in individuals with CHDMs or CHIP.

A limitation of our study is the limited sample size of our cohort. This prevented us from studying the effects of individual genes harboring clonal hematopoiesis driver mutations. A second potential limitation is the sequencing approach utilized in identification of CHDMs. While the use of single-molecule molecular inversion probes allowed for ultrasensitive detection of small clones, the probes were designed against the majority of well-known CH hotspots, except for *DNMT3A* which was covered entirely. Thus, we cannot exclude the possibility that unknown drivers may be located outside the targets included in our assay.

A significant advantage of our study is that we performed extensive phenotyping of metabolic and atherosclerotic clinical parameters, as well as inflammatory and immune parameters. To the best of our knowledge, this study is the first to explore the association between CHDMs and the presence of nonsymptomatic atherosclerotic plaques and immune cell function in overweight and obese individuals. In addition, the use of single-molecule molecular inversion probes allowed us to identify CHDMs with a very low variant allele frequency.

In conclusion, we showed that in overweight and obese individuals, the presence of CHDMs is associated with higher circulating leukocyte and neutrophil numbers, and higher IL-6 concentration, yet with an impaired cytokine production capacity of isolated PBMCs in females. Future studies combining the differential assessment of monocyte and macrophage phenotypes, at both unstimulated and stimulated states are needed. Furthermore, we found no association between CHDMs and the presence of metabolic syndrome or carotid atherosclerotic plaques, supporting the concept that clonal hematopoiesis does not affect atherosclerosis formation *per se*, but might trigger plaque destabilization and the subsequent occurrence of cardiovascular events.

## Data Availability

Part of the data is publically available on the website of Human Functional Genomics Project (HFGP (bbmri.nl)), the rest is available upon reasonable request to the HFGP committee.

## Non-standard Abbreviations and Acronyms

CHIP: Clonal Hematopoiesis of Indeterminate Potential
CHDM: Clonal Hematopoiesis Driver Mutation
VAF: Variant Allele Frequency
PWV: Pulse Wave Velocity
AI: Augmentation Index
NLR: Neutrophil-to-lymphocyte ratio
DNMT3A: DNA methyltransferase 3a
TET2: tet methylcytosine dioxygenase 2

## Acknowledgements

The authors would like to thank the investigators (Inge CL van den Munckhof, Rob ter Horst, Kiki Schraa, Martin Jaeger, Heidi Lemmers and Helga Dijkstra) who performed the inclusion of the study participants and executed laboratory work.

## Supplementary material: Table of contents

**Supplementary table 1:**
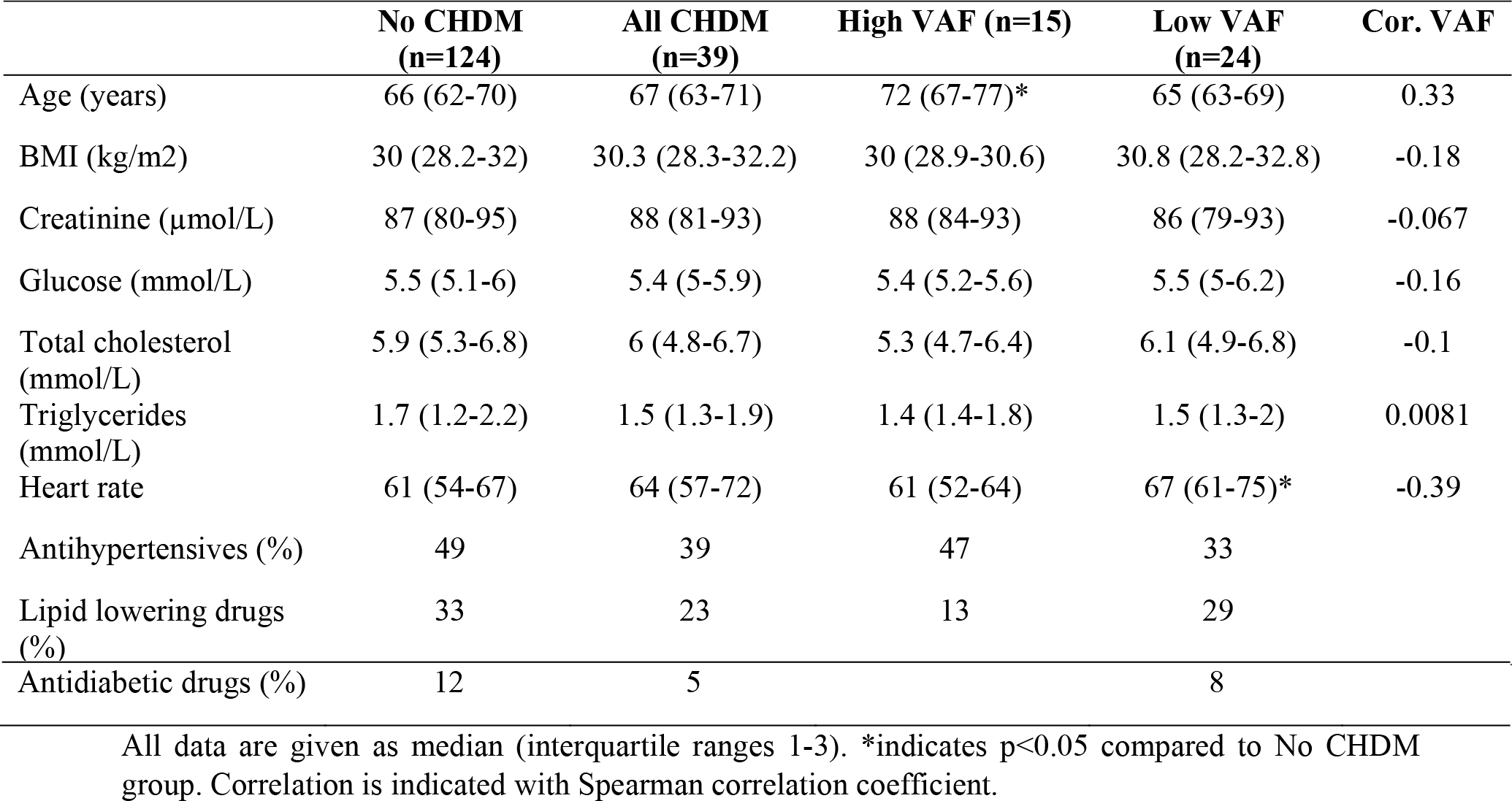
Baseline characteristics of men separated according to CHDM status

**Supplementary table 2:** Baseline characteristics of women separated according to CHDM status

|  | No CHDM<br>(n=88) | All CHDM (n=46) | High VAF (n=18) | Low VAF (n=28) | Cor. VAF |
| --- | --- | --- | --- | --- | --- |
| Age (years) | 67 (64-71) | 68 (63-74) | 64 (63-71) | 68 (65-74) | -0.22 |
| BMI (kg/m<sup>2</sup>) | 30 (28.6-31.9) | 29.6 (28.2-31.9) | 30.4 (28-32.5) | 29.4 (28.4-31.2) | 0.1 |
| Creatinine (μmol/L) | 67 (63-76) | 70 (63-76) | 70 (65-72) | 72 (61-80) | -0.21 |
| Glucose (mmol/L) | 5.4 (5-5.9) | 5.4 (4.9-6) | 5.4 (5-6.3) | 5.2 (5-6) | 0.2 |
| Total cholesterol (mmol/L) | 6.6 (6-7.3) | 6.4 (5.7-7) | 6.7 (5.8-6.9) | 6.4 (5.8-7.2) | -0.1 |
| Triglycerides (mmol/L) | 1.6 (1.3-2.1) | 1.5 (1.3-2.2) | 1.5 (1.3-2.1) | 1.5 (1.3-2.3) | -0.087 |
| Heart rate | 62 (57-68) | 63.5 (59-69) | 65 (59-72) | 63 (53-67) | 0.025 |
| Antihypertensives (%) | 44 | 41 | 56 | 32 |  |
| Lipid lowering drugs (%) | 21 | 26 | 22 | 29 |  |
| Antidiabetic drugs (%) | 6 | 9 | 11 | 7 |  |

All data are given as median (interquartile ranges 1-3). \*indicates $p < 0.05$ compared to No CHDM group. Correlation is indicated with Spearman correlation coefficient.

**Supplementary table 3:**
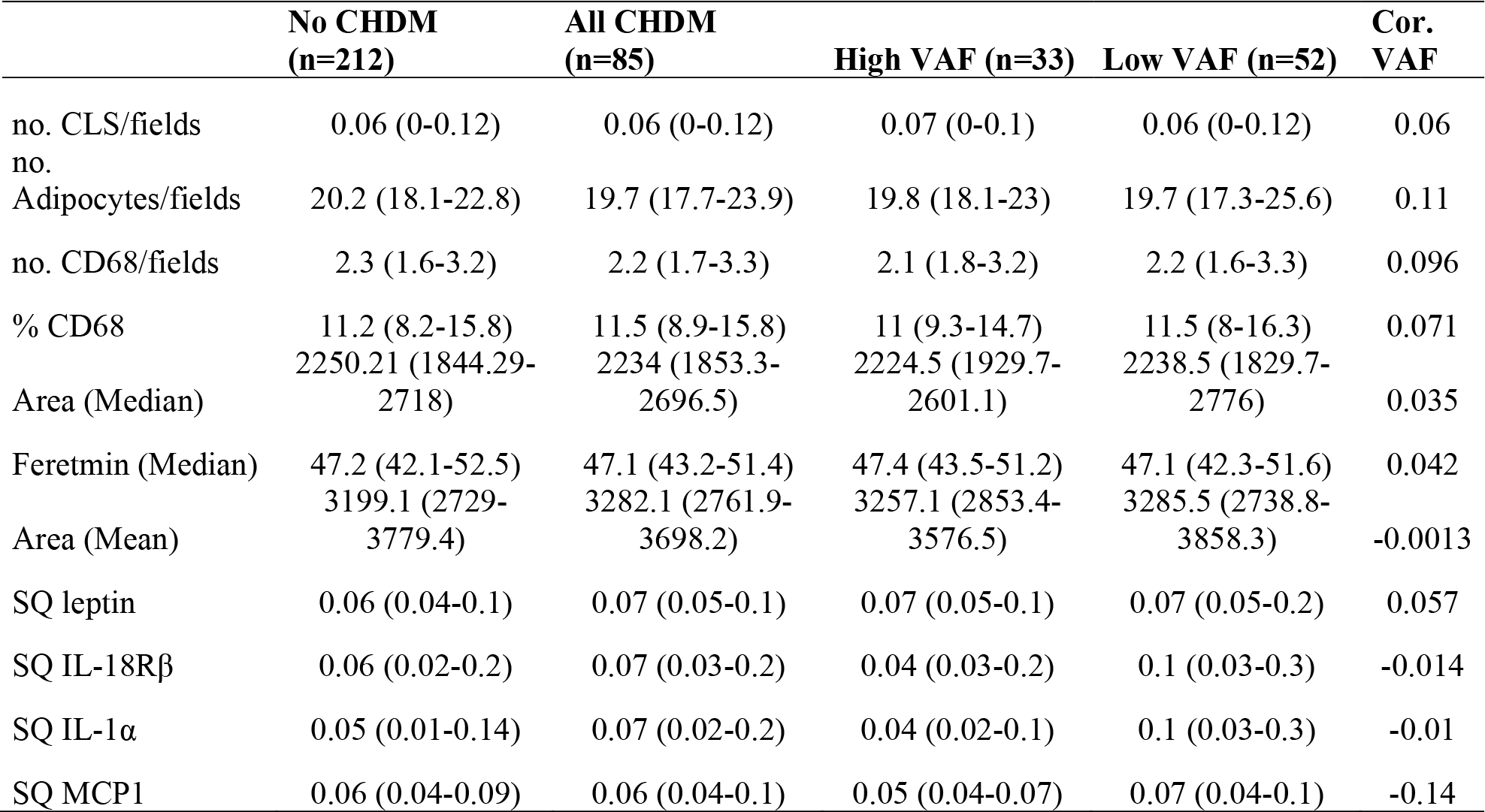
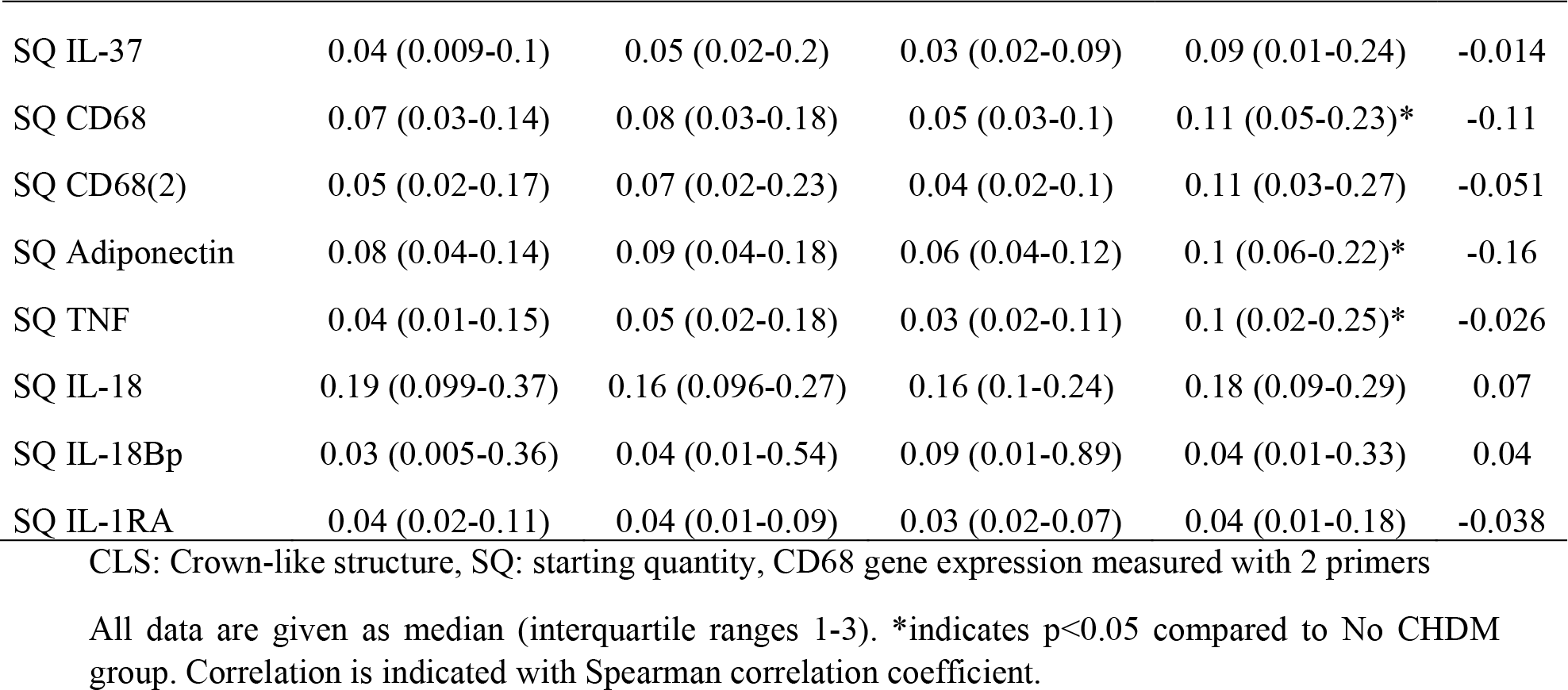
The association between adipose tissue inflammation and CHDM status of the entire study cohort

**Supplementary table 4:** Leukocyte numbers and differentiation, and thrombocyte numbers according to CHDM status of men

|  | No CHDM<br>(n=124) | All CHDM<br>(n=39) | High VAF<br>(n=15) | Low VAF<br>(n=24) | Cor. VAF |
| --- | --- | --- | --- | --- | --- |
| Leukocytes $10^9/l$ | 6 (5-6.7) | 6.3 (5.7-7.4)* | 6.4 (5.9-7.5) | 6.2 (5.6-7.4) | 0.18 |
| Neutrophils $10^9/l$ | 3.2 (2.7-3.8) | 3.6 (2.9-4.8)* | 4 (3.4-4.7)* | 3.4 (2.7-4.7) | 0.2 |
| Lymphocytes $10^9/l$ | 1.8 (1.5-2.2) | 1.8 (1.6-2.1) | 1.7 (1.4-2) | 1.9 (1.6-2.3) | 0.013 |
| Monocytes $10^9/l$ | 0.5 (0.4-0.6) | 0.6 (0.5-0.7) | 0.6 (0.5-0.7) | 0.5 (0.5-0.7) | 0.028 |
| Eosinophils $10^9/l$ | 0.2 (0.1-0.2) | 0.2 (0.1-0.3) | 0.1 (0.1-0.3) | 0.2 (0.1-0.3) | -0.02 |
| Basophils $10^9/l$ | 0.03<br>(0.02-0.04) | 0.03<br>(0.03-0.05)* | 0.03<br>(0.03-0.05)* | 0.03<br>(0.02-0.04) | 0.07 |
| Thrombocytes $10^9/l$ | 217 (188-244) | 208 (193-254) | 215 (199-272) | 206 (190-246) | 0.31 |
| NLR | 1.8 (1.4-2.3) | 2 (1.6-2.5) | 2.3 (1.9-2.7)* | 1.8 (1.2-2.4) | 0.18 |

NLR: Neutrophil to lymphocyte ratio
All data are given as median (interquartile ranges 1-3). \*indicates $p < 0.05$ compared to No CHDM group. Correlation is indicated with Spearman correlation coefficient.

**Supplementary table 5:** Leukocyte numbers and differentiation, and thrombocyte numbers according to CHDM status of women

|  | No CHDM<br>(n=88) | All CHDM<br>(n=46) | High VAF<br>(n=18) | Low VAF<br>(n=28) | Cor. VAF |
| --- | --- | --- | --- | --- | --- |
| Leukocytes 10<sup>9</sup>/l | 5.5 (5-6.3) | 5.9 (5-7.2) | 6.2 (5-7.3) | 5.8 (5.1-6.9) | 0.18 |
| Neutrophils 10<sup>9</sup>/l | 3.1 (2.6-3.6) | 3.2 (2.6-3.9) | 3.2 (2.5-3.9) | 3.1 (2.7-3.9) | 0.14 |
| Lymphocytes 10<sup>9</sup>/l | 1.9 (1.5-2.3) | 2 (1.6-2.3) | 2.1 (1.7-2.3) | 1.9 (1.5-2.3) | 0.2 |
| Monocytes 10<sup>9</sup>/l | 0.4 (0.4-0.5) | 0.5 (0.4-0.6) | 0.5 (0.4-0.6) | 0.4 (0.4-0.5) | 0.27 |
| Eosinophils 10<sup>9</sup>/l | 0.1 (0.1-0.2) | 0.2 (0.1-0.3) | 0.2 (0.1-0.3) | 0.2 (0.1-0.2) | 0.035 |
| Basophils 10<sup>9</sup>/l | 0.03 (0.02-0.04) | 0.03 (0.02-0.04) | 0.03 (0.02-0.04) | 0.03 (0.02-0.04) | -0.046 |
| Thrombocytes 10<sup>9</sup>/l | 247 (216-283) | 247 (215-275) | 247 (205-280) | 248 (226-272) | -0.024 |
| NLR | 1.5 (1.3-2) | 1.7 (1.3-2) | 1.5 (1.3-1.8) | 1.8 (1.4-2) | -0.13 |

NLR: Neutrophil to lymphocyte ratio
All data are given as median (interquartile ranges 1-3). \*indicates p<0.05 compared to No CHDM group. Correlation is indicated with Spearman correlation coefficient.

**Supplementary table 6:** *Ex vivo* cytokine production capacity of PBMCs separated according to CHDM status of men

|  | No CHDM<br>(n=124) | All CHDM (n=39) | High VAF (n=15) | Low VAF (n=24) | Cor.<br>VAF |
| --- | --- | --- | --- | --- | --- |
| LPS(10 ng/ml) | 1029.7 | 1116.9 | 1258.9 | 1099.07 (597.29- |  |
| IL-1β | (543.3-1732) | (589-1657.3) | (602.1-1637.) | 1630.78) | -0.0086 |
| LPS(100 ng/ml) | 2351.1 | 2809.6 | 2894.8 | 2491.5 |  |
| IL-1β | (1475.1-3770) | (1554.1-4038.1) | (2320.3-4038.1) | (1466.8-4080.9) | -0.024 |
|  | 280.3 | 291 | 287 | 315 |  |
| Pam3Cys IL-1β | (112.6-630.6) | (113.9-597.3) | (79-647) | (135.1-475.9) | 0.023 |
| LPS(10 ng/ml) | 5367.8 | 5441.3 | 5342.8 | 6014.2 |  |
| IL-6 | (2940-8598.88) | (4307.7-7309.4) | (4307.65-6801.7) | (4204.9-7363) | -0.18 |
| LPS(100 ng/ml) | 8103.6 | 8383.8 | 7827.2 | 8455.7 |  |
| IL-6 | (4800.8-12236.7) | (5730.1-12896.95) | (5580.3-13232.5) | (6762-12011.8) | -0.14 |
|  | 3750.2 | 3654.1 | 3381.5 | 3744.9 |  |
| Pam3Cys IL-6 | (1590.5-6675.8) | (1602.85-5778.6) | (1100.6-6721.7) | (1998.4-5465.7) | -0.13 |

LPS: Lipopolysaccharide, IL: interleukin, IL-1β and IL-6 concentration units are given in pg/ml
All data are given as median (interquartile ranges 1-3). \*indicates p<0.05 compared to No CHDM group. Correlation is indicated with Spearman correlation coefficient.

**Supplementary table 7:** *Ex vivo* cytokine production capacity of PBMCs separated according to CHDM status of women

|  | No CHDM (n=88) | All CHDM (n=46) | High VAF (n=18) | Low VAF (n=28) | Cor. VAF |
| --- | --- | --- | --- | --- | --- |
| LPS(10 ng/ml) | 1194.1 | 832.2 | 957.3 | 790.1 | 0.12 |
| IL-1β | (677.5-2056.9) | (515.1-1682.4) | (450.4-2448.2) | (533.9-1306.8) | |
| LPS(100 ng/ml) | 2548.5 | 1823.3 | 2054.4 | 1724.3 | 0.11 |
| IL-1β | (1575.3-3638.6) | (1103.3-2950.5) * | (992.7-3010.1) | (1231.2-2833.3) | |
|  | 291.4 | 127.04 | 135.4 | 104.3 | 0.093 |
| Pam3Cys IL-1β | (110.6-671.6) | (64.9-309.92) * | (55.9-397.1) | (67.9-277.2)* | |
| LPS(10 ng/ml) | 6107.8 | 4407.8 | 4407.8 | 4627 | 0.098 |
| IL-6 | (3837.8-9388.1) | (2468-5910.9)* | (2616.3-6051.7)* | (2444.8-5766.8)* |  |
| LPS(100 ng/ml) | 8344.9 | 6349.7 | 5121.7 | 6921.65 | 0.041 |
| IL-6 | (5445.7-13132.3) | (3609.9-8306.8) | (3502-7412)* | (4114.38-10786.5) |  |
|  | 4185.7 | 1569.8 | 1670.3 | 1569.7 | 0.25 |
| Pam3Cys IL-6 | (2125.9-6202.9) | (811.1-2579.2)* | (684.9-3184.6)* | (828-2256.4)* |  |

LPS: Lipopolysaccharide, IL: interleukin, IL-1β and IL-6 concentration units are given in pg/ml
All data are given as median (interquartile ranges 1-3). \*indicates p<0.05 compared to No CHDM group. Correlation is indicated with Spearman correlation coefficient

**Supplementary table 8:** Circulating cytokines and adipokines in plasma separated according to CHDM status of the men

|  | No CHDM (n=124) | All CHDM (n=39) | High VAF (n=24) | Low VAF (n=15) | Cor. VAF |
| --- | --- | --- | --- | --- | --- |
| IL-6 (pg/ml) | 2.3 (1.6-3.3) | 2.8 (1.8-4.6) | 2.9 (2.1-4.6) | 2.7 (1.7-4) | 0.052 |
| IL-1β (pg/ml) | 0.06 (0.06-0.1) | 0.06 (0.06-0.11) | 0.06 (0.06-0.01) | 0.08 (0.06-0.11) | -0.1 |
| IL-18 (pg/ml) | 313 (222.7-515) | 300.9 (224.5-510.2) | 277.7 (224.5-349.2) | 341 (232.5-628.4) | -0.17 |
| IL-18bpa (ng/ml) | 17.1 (14.4-20) | 16.4 (13.6-20.2) | 16.5 (13-17.7) | 16.2 (14.1-21.2) | -0.2 |
| hsCRP (μg/ml) | 1.4 (0.9-3) | 1.9 (1.1-3.1) | 1.9 (1-3.7) | 1.9 (1.3-2.8) | 0.033 |
| AAT (mg/ml) | 1 (0.6-1.7) | 0.9 (0.6-1.4) | 0.9 (0.7-1.1) | 0.87 (0.61-1.44) | 0.12 |
| Resistin (ng/ml) | 10.3 (8-12.8) | 11.5 (9.3-15.2) | 11.8 (10-14.9) | 11.3 (9.2-15.9) | -0.0066 |
| Leptin (ng/ml) | 10.2 (7-15) | 12.3 (7.5-17) | 8.5 (6.9-16.1) | 13.3 (8.4-21.3) | -0.22 |
| Adiponectin (μg/ml) | 3.1 (2.2-4.5) | 3.3 (2.7-4.3) | 3.7 (3-4.2) | 3.1 (2.3-4.4) | 0.092 |

CHDM status of men
IL: interleukin, IL-18bp: IL-18 binding protein, CRP: C-reactive protein, AAT: Alpha-1 antitrypsin
All data are given as median (interquartile ranges 1-3). \*indicates $p < 0.05$ compared to No CHDM group. Correlation is indicated with Spearman correlation coefficient.

**Supplementary table 9:** Circulating cytokines and adipokines in plasma separated according to CHDM status of women

|  | No CHDM (n=88) | All CHDM (n=46) | High VAF (n=18) | Low VAF (n=28) | Cor. VAF |
| --- | --- | --- | --- | --- | --- |
| IL-6 (pg/ml) | 2.7 (1.9-3.4) | 2.6 (1.7-4.1) | 3.3 (2-4.4) | 2.2 (1.5-3) | 0.36 |
| IL-1β (pg/ml) | 0.06 (0.06-0.11) | 0.06 (0.06-0.13) | 0.06 (0.06-0.14) | 0.07 (0.06-0.11) | 0.013 |
| IL-18 (pg/ml) | 300.1 (234.1-478.8) | 283.7 (204.2-469.9) | 411.3 (222.3-489.9) | 252.5 (200-379) | 0.27 |
| IL-18bpa (ng/ml) | 17.5 (14-21.4) | 15.8 (13.5-19) | 14.9 (12.8-18.7) | 16.2 (14.2-19.2) | -0.098 |
| hsCRP (μg/ml) | 2.7 (1.1-4.6) | 2.1 (1.3-3.7) | 2 (0.9-3.7) | 2.2 (1.5-3.8) | 0.13 |
| AAT (mg/ml) | 0.9 (0.6-1.9) | 0.9 (0.6-1.3) | 0.8 (0.6-1.6) | 0.9 (0.6-1.1) | -0.037 |
| Resistin (ng/ml) | 10.8 (8.6-15.3) | 11.5 (9-14.4) | 11.4 (8.3-13) | 12.8 (10-16.2) | -0.18 |
| Leptin (ng/ml) | 31.5 (21.7-48.1) | 27.5 (17.8-51) | 25.4 (18.8-47.1) | 28.8 (17.5-52.7) | -0.056 |
| Adiponectin (μg/ml) | 6 (4.6-7.3) | 5.1 (4.2-8) | 4.9 (4.2-6.8) | 6 (4.2-8) | -0.046 |

IL: interleukin, IL-18bp: IL-18 binding protein, CRP: C-reactive protein, AAT: Alpha-1 antitrypsin
All data are given as median (interquartile ranges 1-3). \*indicates $p < 0.05$ compared to No CHDM group. Correlation is indicated with Spearman correlation coefficient.

**Supplementary table 10:** The association between targeted proteomic biomarkers and CHDM status of the entire study cohort

|  | No CHDM (n=212) | All CHDM (n=85) | High VAF (n=33) | Low VAF (n=52) | Cor. VAF |
| --- | --- | --- | --- | --- | --- |
| SLAMF7 | 3.6 (3.3-4.1) | 3.5 (3.2-3.9) | 3.6 (3.3-4) | 3.4 (3-3.7)* | 0.22 |
| IL1RL2 | 4.5 (4.3-4.8) | 4.6 (4.4-4.9)* | 4.6 (4.3-4.8) | 4.6 (4.4-4.9) | 0.15 |
| IL-27 | 6.5 (6.3-6.7) | 6.4 (6.1-6.7)* | 6.3 (6.1-6.7) | 6.4 (6.1-6.6) | 0.021 |
| GH | 8 (6.5-9.4) | 7.3 (6.3-9.3) | 7.9 (6.3-9.9) | 7.2 (6.2-8.7)* | 0.23 |
| GLO1 | 5.6 (5.4-5.9) | 5.5 (5.1-5.8) | 5.4 (5.1-5.8)* | 5.6 (5.2-5.9) | -0.21 |
| AMBP | 7.9 (7.9-8) | 8 (7.8-8.1) | 7.9 (7.8-8) | 8 (7.9-8.1)* | -0.073 |
| CCL3 | 6.7 (6.3-7) | 6.7 (6.1-7) | 6.4 (6.1-6.7)* | 6.7 (6.4-7.1) | -0.26 |
| TNFRSF13B | 10 (9.8-10.3) | 9.9 (9.7-10.1)* | 10 (9.8-10.2) | 9.9 (9.7-10.1)* | 0.042 |
| LEP | 6.8 (6.2-7.4) | 7.1 (6.5-7.5)* | 7.1 (6.4-7.4) | 7.2 (6.5-7.6)* | 0.012 |
| NEMO | 5.8 (5.2-6.4) | 5.7 (5-6.2) | 5.5 (5-5.8)* | 5.9 (5-6.4) | -0.21 |
| VEGFD | 8.1 (7.9-8.3) | 8 (7.8-8.2) | 8.1 (7.9-8.4) | 8 (7.8-8.1)* | 0.33 |
| AXIN1 | 2.7 (2.3-3.2) | 2.6 (2-3.2) | 2.3 (2-3)* | 2.7 (2.1-3.5) | -0.16 |
| TSLP | 0.6 (0.4-0.8) | 0.6 (0.4-0.8) | 0.5 (0.3-0.7)* | 0.6 (0.4-0.8) | -0.14 |
| MMP-1 | 9.7 (9-10.4) | 9.6 (9-10.2) | 9.4 (8.9-9.7)* | 9.8 (9.2-10.5) | -0.23 |
| CCL3.1 | 5.1 (4.9-5.5) | 5.1 (4.8-5.4) | 4.9 (4.8-5.2)* | 5.2 (4.8-5.7) | -0.22 |
| FGF-19 | 8.3 (7.7-8.9) | 8 (7.4-8.5)* | 8.1 (7.5-8.6) | 7.9 (7.3-8.5)* | 0.081 |
| ST1A1 | 2.9 (2.3-3.5) | 2.8 (2.1-3.4) | 2.4 (2-3.1)* | 2.9 (2.1-3.5) | -0.17 |

All data are given as median (interquartile ranges 1-3). \*indicates $p < 0.05$ compared to No CHDM group. Correlation is indicated with Spearman correlation coefficient.

**Supplementary table 11 (Excel):** List of 24 genes sequenced and known clonal hematopoiesis hotspots

**Supplementary table 12 (Excel):** single-molecule Molecular Inversion Probes (smMIP)

**Supplementary table 13 (Excel):** Variant characteristics of candidate clonal hematopoiesis driver mutations in this study

